# Delirium due to hip fracture is associated with activated immune-inflammatory pathways and a reduction in negative immunoregulatory mechanisms

**DOI:** 10.1101/2022.02.28.22271663

**Authors:** Paul Thisayakorn, Yanin Thipakorn, Saran Tantavisut, Sunee Sirivichayakul, Michael Maes

**Author notes:** Corresponding authors: Dr. Paul Thisayakorn, M.D., Prof. Dr. Michael Maes, M.D., Ph.D., Department of Psychiatry, Faculty of Medicine, Chulalongkorn University, Bangkok, Thailand, <u>(14) Michael Maes | Stats (researchgate.net)</u>, <u>Michael Maes - Google Scholar</u>.

## Abstract

The objectives of this study were to delineate whether delirium is associated with activation of the immune-inflammatory response system (IRS) as indicated by activation of M1, Thelper (Th)1, and Th17 profiles, and/or by reduced activities of the compensatory immunoregulatory system (CIRS), including Th2 and Tregulatory profiles. We recruited 65 elderly patients with a low energy impact hip fracture who underwent hip fracture operation. The CAM-ICU and the Delirium Rating Scale, Revised-98-Thai version (DRS-R-98) were assessed pre-operatively and 1, 2 and 3 days after surgery. Blood samples (day 1 and 2) post-surgery were assayed for cytokines/chemokines using a MultiPlex assay and the neutrophil/lymphocyte ratio. We found that delirium and/or the DRS-R-98 score were associated with IRS activation as indicated by activated M1, Th1, Th17 and T cell growth profiles and by attenuated CIRS functions. The most important IRS biomarkers were CXCL8, interleukin (IL)-6, and tumor necrosis factor-α, and the most important CIRS biomarkers were IL-4 and soluble IL-1 receptor antagonist. We found that 42.5% of the variance in the actual changes in the DRS-R-98 score (averaged from day 1 to day 3) was explained by T cell growth factors, baseline DRS-R-98 scores and age. The pain scores during delirium were significantly and positively associated with CXCL8 and CCL3 and negatively with IL-4 and sIL-1RA. An increase in the NLR reflects overall IRS, M1, Th1, Th17, and Th2 activation. In conclusion, post-hip surgery delirium is associated with activated IRS pathways and appears especially in patients with lowered CIRS functions.

## Introduction

Delirium is a neuropsychiatric syndrome presenting with altered levels of attention, awareness, and cognitive functions [1, 2]. Delirium is commonly observed in hospitalized elderly individuals who require surgical management for hip fracture surgery [3-5] with a prevalence of pre-operative [6, 7] and post-operative hip fracture delirium of 12.7-57.6% and 16.9-24%, respectively [8, 9]. Increased risk of medical comorbidities, prolonged length of intensive care unit and hospital stay, as well as higher mortality are documented in hip fracture patients with delirium [10-12].

During the acute period of severe physical illness, delirium may develop as the final pathway of intertwined systemic and central nervous system pathogenic processes [13, 14]. The aging brain, circadian rhythm disturbances, stress responses including in the endocrine system, increased oxidative stress, neurotransmitter dysregulations, neuronal circuit disruptions, as well as activation of immune-inflammatory pathways contribute to the pathophysiology of delirium [15, 16]. We found a significant association between the onset of delirium following hip fracture and increased white blood cell number, neutrophil percentage, neutrophil / lymphocyte ratio (NLR), and blood gas parameters including elevated pO2 values [17]. The most significant biomarker of delirium was an increase in the NLR [17], indicating that delirium due to hip fracture may be caused by an aseptic immune-inflammatory process originating from hip tissue trauma which triggers a more generalized immune response. In this respect, previous reports showed that increases in peripheral levels of C-reactive protein (CRP), interleukin-6 (IL-6), CXCL8 (IL-8), and tumor necrotic factor (TNF)-α are associated with the onset of post-operative delirium [3, 18-20]. However, not all research came to the same conclusion all of the time. For example, one study showed increased IL-6, IL-2, and IL-1α levels in delirium due to septic shock, although no such increases in CRP, CXCL8 and TNF-α could be established [21].

Overall, the findings reflect peripheral activation of immune-inflammatory pathways which is triggered by local tissue injury and/or surgical damage ultimately leading to neuro-inflammatory signaling in the brain which may contribute to the symptoms of delirium [22]. It is interesting to note that mild chronic activation of immune-inflammatory pathways occurs in schizophrenia [23, 24], affective disorders [25, 26], and dementia [27], which are known risk factors of delirium [28]. Moreover, increased neurotoxicity due to the cumulative effects of M1 macrophage, T helper (Th)1, and Th17 phenotypes and neurotoxic cytokines/chemokines e.g., CCL11, CCL2, RANTES (CCL5), CXCL10 (IP-10) and CCL3 (macrophage inflammatory protein 1α) to a large extent explain the symptoms and cognitive impairments of the major psychoses [24, 29, 30] and, therefore, could be involved in the pathophysiology of delirium.

Moreover, the compensatory immune regulatory system (CIRS), which may attenuate an overzealous inflammatory response, is activated in affective disorders and schizophrenia and these include T regulatory (Treg) (e.g., IL-10) and Th2 (e.g., IL-4, IL-9, and IL-13) profiles. However, there are no data whether delirium is associated with M1 macrophage, Th1, Th2, Th17, Treg or CIRS cytokine profiles, and whether a neurotoxic cytokine profile including M1, Th1, Th17 and neurotoxic chemokines, such as CCL11, CCL2, CCL3, CCL5, and CXCL10) is associated with delirium.

Complex interaction between pain and delirium especially in elderly demented people are frequently described [31]. Pain may increase the risk of developing delirium in both general hospital and skilled nursing facilities [32, 33]. Furthermore, post-injury associated peripheral and central pain cascades may be mediated by increased levels of selected proinflammatory cytokines and chemokines such as IL-1β, IL-6, TNF-α, CCL2, CCL3, and CCL5 [34, 35]. Similar peripheral immune aberrations were also observed in chronic pain conditions as shown by a recent meta-analysis reporting that IL-6, IL-4, IL-17, and M1 macrophages are upregulated in fibromyalgia patients as compared with healthy controls [36]. Nevertheless, there are no data whether delirium is accompanied by higher pain scores in association with increased levels of proinflammatory cytokines and chemokines.

Hence, the aims of this study were to delineate a) the cytokine profiles (including M1 macrophage, Th1, Th2, Th17, Treg, Tcell growth) of delirium due to hip fracture; b) whether a neurotoxic cytokine/chemokine profile consisting of M1, Th1, Th17 cytokines and the neurotoxic CCL11, CCL2, CCL3, CCL5,CXCL8 and CXCL10 chemokines are associated with delirium; and c) whether increases in these immune markers are also associated with increased pain scores. The specific hypotheses are that a) delirium is characterized by increased M1, Th1, and Th17-neurotoxicity profiles and lowered CIRS, Treg and Th2 profiles, and b) pro-inflammatory cytokines are associated with increased pain levels. Moreover, since the NLR is a major biomarker of delirium we also examined the cytokine profiles, which are associated with the increased NLR.

## Subjects and Methods

### Participants

We recruited sixty-five elderly hip fracture patients who were admitted into the Hip Fracture Pathway Inpatient Care at King Chulalongkorn Memorial Hospital, Bangkok, Thailand between June, 2019 and February, 2020. Patients aged 65-year and older and who suffered from a low energy impact hip fracture and underwent a hip fracture operation and were postoperatively transferred to the surgery intensive care unit (SICU) or orthopedic units were included into the study. The diagnosis of delirium was made using the Confusion Assessment Method-Intensive Care Unit-Thai version (CAM-ICU) [37] and DSM-5 criteria of delirium [1]. Exclusion criteria were: a high energy impact hip fracture, metastatic fractures, intracranial vascular lesion and other traumatic brain injury from falling, coma or premorbid dementia, a life-time history of (neuro)-inflammatory and/or neurodegenerative disease including multiple sclerosis, Parkinson’s and Alzheimer’s disease, rheumatoid arthritis, inflammatory bowel disease, and major psychiatric illness such as schizophrenia, bipolar disorder, and the acute phase of a major depressive disorder. Remitted patients with major depression, stroke patients one year after the acute stroke and subjects with mild cognitive impairment could be included. We also excluded patients who could not communicate in Thai language.

### Clinical assessments

Initially, the demographic and clinical information of the research participants was extracted from the electronic medical records and bedside interviews. The baseline cognitive status and delirium scores were assessed within 24 hours before the surgery date. Then, we re-assessed daily the cognitive status and delirium severity and diagnosis during three consecutive days postoperatively. The CAM-ICU and Delirium Rating Scale, Revised-98-Thai version (DRS-R-98) were used at bedside to determine the presentation and the severity of the delirium in the evening of day 0 (pre-operative day), and the three consecutive days after the surgery. Both CAM-ICU-T and DRS-R-98-T show good sensitivity, specificity for delirium and interrater reliability [37, 38]. The nursing staff monitored the post-operative pain by using a 1-10 visual analog pain scale. We recorded the use of anticholinergic medications, benzodiazepines, opiates, and psychotropic drugs prior to hospitalization, as well as pertinent peri/post-operative clinical data, such as operative time, blood loss, and the need for restraint due to psychomotor agitation.

The Faculty of Medicine, Chulalongkorn University, Bangkok, Thailand (registration number 528/61) institutional review board reviewed and approved this study in accordance with the International Guideline for the Protection of Human Subjects, as required by the Declaration of Helsinki, The Belmont Report, CIOMS Guideline, and International Conference on Harmonization in Good Clinical Practice (ICH-GCP). All patients and their guardians (first degree family members) provided written informed consent.

### Assays

Along with the clinical evaluation, venous blood samples were collected post-surgery, daily at 7.00 am, for two consecutive days. Blood samples were sent to the laboratory to assay complete blood counts and venous blood gas. Plasma and serum samples were frozen at -80°C until thawed for the assay of cytokines/chemokines. The CBC (NLR) and blood gas (HCO3-) assays were performed as described previously [17]. The CBC values were determined using a flow cytometry method with a semiconductor laser (Sysmex, Kobe, Japan) and the NLR were determined from the CBC results. The NLR was calculated as a z unit-based composite score equal to the difference between the z scores of neutrophil and z scores of lymphocyte percentages. The blood gas data were analyzed using an ion-selective electrode from the Nova Stat Profile pHOx series (Nova Biomedica, MA, USA).

To assay cytokines/chemokines, we used the Bio-Plex Pro™ Human Chemokine Assays (Bio-Rad Laboratories, Inc. USA). We assayed IL-1β, IL-1Ra, IL-2, IL-4, IL-5, IL-6, IL-7, IL-8, IL-9, IL-10, IL-12 (p70), IL-13, IL-15, IL-17, basic FGF, CCL11 (eotaxin), G-CSF, GM-CSF, IFN-γ, CXCL10, CCL2, CCL3, CCL4, PDGF, CCL5, TNF-α, and VEGF. Fifty microlitres of serum (1:4 dilutions in sample diluent HB) was mixed with 50 µl of microparticle cocktail (containing cytokine/chemokines per well of a 96-well plate provided by the manufacturer) and incubated for 1 hour at room temperature while shaking at 850 rpm. Wells were washed three times before another 50 µl of diluted Streptavidin-PE was added and further incubated for 10 minutes at room temperature on shaker at 850 rpm. Finally, wells were washed three times and 125 µl of assay buffer was added and shake at 850 rpm at room temperature for 30 seconds before being read with Bio-Plex^®^ 200 System (Bio-Rad Laboratories, Inc.). In the data analyses we used the concentrations of the cytokines/chemokines. More than 20% of all measured concentrations of IL-2, IL-5, IL-10, IL-12, IL-13, IL-15, GM-CSF and VEGF were below the detection limit and, therefore, these cytokines/growth factors were excluded from the analyses concerning the effects of single cytokines/growth factors. Nevertheless, these values were considered when computing immune profiles because measurable levels of those cytokines/growth factors may contribute to IRS/CIRS/ T cell growth responses. The primary outcome variables in this study were different immune profiles, namely the M1 macrophage profile computed as z IL-1β + z IL-6 + zTNF-α + z CXCL8 + z CCL3 + z sIL-RA; Th1 as z IL-2 + z IL-12 + z IFN (interferon)-γ; Th2: z IL-4 + z IL-9 + z IL-13; Th17: z IL-6 + z IL-17; the IRS/CIRS ratio as z (M1 +Th1+ Th-17) – z (z IL-4 + z IL-9 + z IL-13 + z IL-10); T cell growth (all factors that promote T cell growth): z IL-4 + z IL-7 + z IL-9 + z IL-12 + z IL-15 + z GM-CSF (granulocyte-macrophage colony-stimulating factor). Neurotoxicity was conceptualized as a composite score comprising neurotoxic cytokines/chemokines: z IL-1β + z TNF-α + z IL-6 + z IL-2 + z IFN-γ + z IL-17 + zCCL11 + z CXCL10 + z CCL3 + z CCL5 + z CCL2. The intra-assay CV values for all analytes were < 11.0%.

### Statistics

The X^2^-test was used to determine associations between sets of categorical variables, and analysis of variance (ANOVA) was used to determine between-group differences in scale variables. The primary outcome measures are the delirium diagnosis, as determined by the CAM-ICU and DSM-IV-TR, and the quantitative DRS-R-98 scale scores. The primary statistical analysis used generalized estimating equations (GEE) to examine the associations between the immune profiles and NLR on the outcome (delirium as binary variable), or multiple regression analyses which examined the effects of immune profiles, HCO3- and clinical variables (previous MDD and CNS disease, age, sex, BMI, time to surgery, estimated blood loss during surgery, duration of surgery, use of deliriogenic medications, insomnia, nasal cannula oxygen) on the DSR-R-98 scores while adjusting for the baseline DRS-R-98 scores. The latter regression analysis estimates the effects of biomarkers on the actual changes in DRS-R-98 score from baseline to day 1, 2 or 3. Multiple comparisons among treatment means or multiple associations between outcome data and immune profiles were adjusted using False Discovery Rate (FDR) p-correction. Automatic multivariate regression analysis was employed to predict dependent variables (the DRS-R-98 scores) using immune profiles, NLR and chemokines and demographic data, while examining R^2^ changes, multicollinearity (using tolerance and VIF), multivariate normality (Cook’s distance and leverage), and homoscedasticity (using White and modified Breusch-Pagan tests for homoscedasticity). We used an automatic stepwise (step-up) procedure with a 0.05 p-to-enter and a 0.06 p-to-remove. The results of all these regression analyses were always bootstrapped using 5.000 bootstrap samples, and the latter are shown if the results were not concordant. IBM SPSS Windows version 25, 2017 was used for statistical analysis and statistical significance was set at <0.05 (two tailed tests). Using G power analysis, the sample size for a repeated measurement design ANOVA should be around n = 60 when the effect size is 0.3, alpha is 0.05, power is 0.80, including two groups (delirium versus non-delirium) and two repeated (the biomarkers) measurements. The a priori computed required sample size for a multiple regression analysis given an effect size of 0.3, alpha 0.05, power 0.80, number of predictors five is around forty-nine. Therefore, in the present study sixty-five patients were included.

## Results

A total of sixty-five elderly patients with hip fracture and who underwent orthopedic surgery were included into the study. Nineteen of them (29.2%) developed delirium peri-operatively. **Table 1** shows the socio-demographic and clinical data in both patients with and without delirium. The delirious patients showed a higher mean age and lower body mass index (BMI), and a longer waiting time to surgery. There were no differences in sex ratio, marital status, surgical time, blood loss during surgery and the pain scores between both study groups. There were no significant differences in HCO3- and insomnia between both groups, while the prevalence of CNS disease and stroke was higher in those with a delirium.

**Table 1.** Socio-demographic and clinical data of hip surgery patients divided into those with and without delirium

| Variables | No Delirium (N=46) | Delirium (N=19) | F/X<sup>2</sup> | df | p |
| --- | --- | --- | --- | --- | --- |
| Age (years) | 79.7 (7.9) | 84.5 (6.3) | 5.57 | 1/63 | 0.021 |
| Sex (female/male) | 36/10 | 15/4 | 0.00 | 1 | 0.951 |
| Education (years) | 7.5 (5.6) | 9.4 (6.9) | 1.44 | 1/61 | 0.235 |
| BMI (kg/m<sup>2</sup>) | 22.3 (3.2) | 19.7 (2.7) | 8.42 | 1/57 | 0.005 |
| Marital status (single/married/divorced) | 5/20/1 | 6/6/0 | FEPT | - | 0.233 |
| Time to surgery (Hours) | 60.8 (47.9) | 109.1 (101.5) | 6.79 | 1/62 | 0.011 |
| Surgical time (Minutes) | 104.5 (42.8) | 97.9 (29.9) | 0.38 | 1/63 | 0.543 |
| Blood loss (mL) | 199.8 (110.8) | 231.6 (138.7) | 0.95 | 1/63 | 0.333 |
| Pain score day 0-1 | 2.57 (2.09) | 3.44 (2.31) | 2.14 | 1/61 | 0.149 |
| Pain score day 2-3 | 2.67 (2.69) | 2.17 (2.18) | 0.49 | 1/61 | 0.486 |
| HCO<sub>3</sub><sup>-</sup> mEq/L | 27.1 (3.4) | 26.8 (3.5) | 0.09 | 1/63 | 0.762 |
| CNS disease (N/Y) | 36/10 | 7/12 | 10.30 | 1 | 0.002 |
| Insomnia (N/Y) | 41/5 | 17/2 | 0.00 | 1 | 0.968 |
| Stroke (N/Y) | 42/4 | 12/7 | 7.58 | 1 | 0.006 |
| Depression (N/Y) | 42/4 | 18/1 | FEPT | - | 1.000 |

Results are shown as mean ± SD. All results of analysis of variance (F values), analysis of contingency analysis (X<sup>2</sup>) or Fisher exact probability test (FEPT)
BMI: body mass index

**Table 2** shows the results of GEE analysis performed with delirium as dependent variables and the immune profiles and NLR as explanatory variables. The changes in the DRS-R-98 scores from baseline to the mean values at day 1, 2 and 3 were significantly higher in delirium patients as compared with those without delirium. After FDR p-correction, the IRS/CIRS ratio, M1, Th17, T cell growth, and NLR were significantly and positively associated with delirium (no delirium as reference group).

**Table 2.**
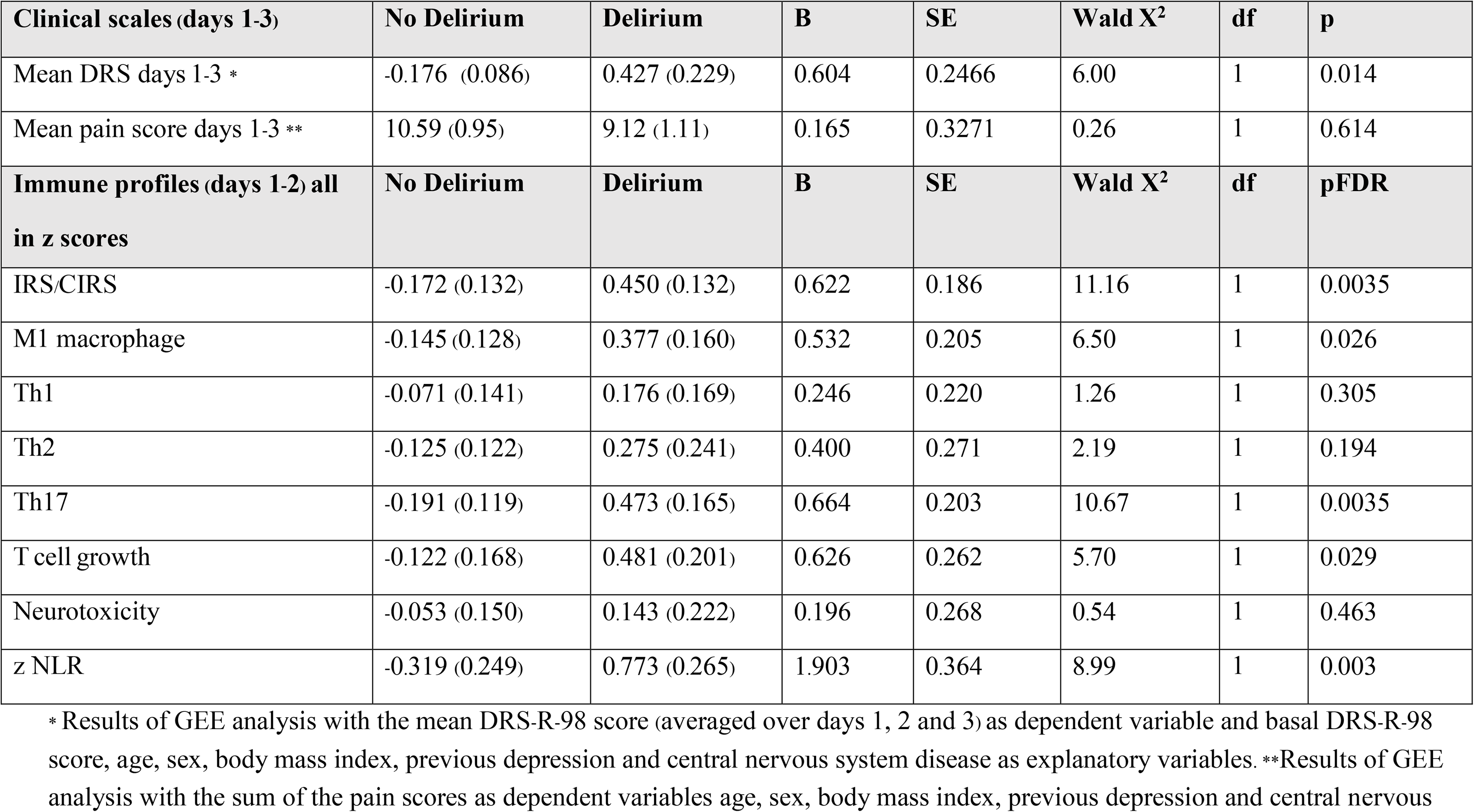
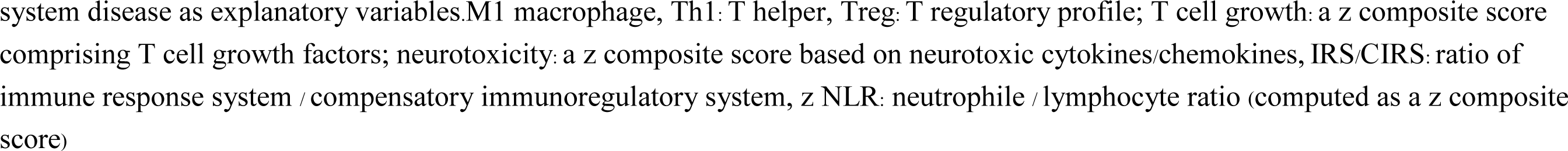
Differences in Delirium Rating Scale, Revised-98-Thai (DRS) and pain scores and immune profiles between patients with and without delirium

| **Clinical scales (days 1-3)** | **No Delirium** | **Delirium** | **B** | **SE** | **Wald X<sup>2</sup>** | **df** | **p** |
| --- | --- | --- | --- | --- | --- | --- | --- |
| Mean DRS days 1-3 * | -0.176 (0.086) | 0.427 (0.229) | 0.604 | 0.2466 | 6.00 | 1 | 0.014 |
| Mean pain score days 1-3 ** | 10.59 (0.95) | 9.12 (1.11) | 0.165 | 0.3271 | 0.26 | 1 | 0.614 |
| **Immune profiles (days 1-2) all in z scores** | **No Delirium** | **Delirium** | **B** | **SE** | **Wald X<sup>2</sup>** | **df** | **pFDR** |
| IRS/CIRS | -0.172 (0.132) | 0.450 (0.132) | 0.622 | 0.186 | 11.16 | 1 | 0.0035 |
| M1 macrophage | -0.145 (0.128) | 0.377 (0.160) | 0.532 | 0.205 | 6.50 | 1 | 0.026 |
| Th1 | -0.071 (0.141) | 0.176 (0.169) | 0.246 | 0.220 | 1.26 | 1 | 0.305 |
| Th2 | -0.125 (0.122) | 0.275 (0.241) | 0.400 | 0.271 | 2.19 | 1 | 0.194 |
| Th17 | -0.191 (0.119) | 0.473 (0.165) | 0.664 | 0.203 | 10.67 | 1 | 0.0035 |
| T cell growth | -0.122 (0.168) | 0.481 (0.201) | 0.626 | 0.262 | 5.70 | 1 | 0.029 |
| Neurotoxicity | -0.053 (0.150) | 0.143 (0.222) | 0.196 | 0.268 | 0.54 | 1 | 0.463 |
| z NLR | -0.319 (0.249) | 0.773 (0.265) | 1.903 | 0.364 | 8.99 | 1 | 0.003 |

\* Results of GEE analysis with the mean DRS-R-98 score (averaged over days 1, 2 and 3) as dependent variable and basal DRS-R-98 score, age, sex, body mass index, previous depression and central nervous system disease as explanatory variables. \*\*Results of GEE analysis with the sum of the pain scores as dependent variables age, sex, body mass index, previous depression and central nervous
system disease as explanatory variables. M1 macrophage, Th1: T helper, Treg: T regulatory profile; T cell growth: a z composite score comprising T cell growth factors; neurotoxicity: a z composite score based on neurotoxic cytokines/chemokines, IRS/CIRS: ratio of immune response system / compensatory immunoregulatory system, z NLR: neutrophil / lymphocyte ratio (computed as a z composite score)

**Table 3** shows the results of multiple regression analyses with the DRS-R-98 scores at day 1, day 2 and day 3 as dependent variables and cytokine profiles as explanatory variables while allowing for the effects of demographic/clinical data. Regression # 1 shows that 33.0% of the variance in the DRS-R-98 score at day 1 was explained by CNS disease and IL-8 (day 1) (both positively associated) and BMI (inversely associated). Regression # 2 shows that 31.3% of the variance in the DRS-R-98 score at day 2 was explained by the cumulative effects of T cell growth profile (positively associated) and BMI and IL-4 (both inversely). **Figure 1** shows the partial regression of the DRS-R-98 score (day 2) on the T cell growth profile measured at day 1 after controlling for the variables shown in regression # 2. **Figure 2** shows the partial regression of the DRS-R-98 score (day 2) on IL-4 (day 1). Regression # 3 shows that 35.5% of the variance in the DRS-R-98 score (day 2) was explained by a combination of Th1 profile (day 2), age, CNS disorders (positively), and sIL-1RA (day 2 an inversely). Regression # 4 shows that 15.9% of the variance in the DRS-R-98 score (day 3) was positively associated with the IRS/CIRS ratio at day 2. **Figure 3** shows the partial regression of the DRS-R-98 score (day 3) on the IRS/CIRS ratio at day 2 after controlling for age (not significant). Regression #5 shows that 15.3% of the variance in the mean DRS-R-98 score averaged over days 1, 2 and 3 was explained by the regression on IL-6 (day 1 and positively) and BMI (inversely).

**Table 3.** Results of multiple regression analysis with the Delirium Rating Scale, Revised-98-Thai (DRS) scores at day 1, day 2 and day 3 as dependent variables and immune/cytokine profiles as explanatory variables while allowing for the effects of demographic/clinical data.

| Dependent variables | Explanatory variables | B | t | p | F model | df | p | R<sup>2</sup> |
| --- | --- | --- | --- | --- | --- | --- | --- | --- |
| # 1 DRS day 1 | **Model** |  |  |  | **10.03** | **3/61** | **<0.001** | **0.330** |
|  | CNS disease | 0.450 | 4.26 | <0.001 |  |  |  |  |
|  | IL-8 - day 1 | 0.263 | 2.51 | 0.015 |  |  |  |  |
|  | BMI | -0.262 | -2.49 | 0.016 |  |  |  |  |
| #2 DRS day 2 | **Model** |  |  |  | **9.13** | **3/60** | **<0.001** | **0.313** |
|  | BMI | -0.283 | -2.61 | 0.011 |  |  |  |  |
|  | T cell growth - day1 | 0.732 | 4.13 | <0.001 |  |  |  |  |
|  | IL-4 - day 1 | -0.558 | -3.13 | 0.003 |  |  |  |  |
| #3 DRS day 2 | **Model** |  |  |  | **7.84** | **4/57** | **<0.001** | **0.355** |
|  | Th1 - day 2 | 0.397 | 3.54 | <0.001 |  |  |  |  |
|  | Age | 0.370 | 3.19 | 0.002 |  |  |  |  |
|  | sIL-1RA - day 2 | -0.289 | -2.48 | 0.016 |  |  |  |  |
|  | CNS disease | 0.228 | 2.12 | 0.039 |  |  |  |  |
| #4 DRS day 3 | **Model** |  |  |  | **11.88** | **1/63** | **<0.001** | **0.159** |
|  | IRS/CIRS - day 2 | 0.398 | 3.45 | <0.001 |  |  |  |  |
| #5 Mean DRS days 1–3 | **Model** |  |  |  | **5.51** | **2/61** | **0.006** | **0.153** |
|  | IL-6 - day 1 | 0.313 | 2.65 | 0.010 |  |  |  |  |
|  | BMI | -0.244 | -2.07 | 0.043 |  |  |  |  |

CNS: Central nervous system, IL: interleukin, BMI: body mass index, Th: T helper, IRS/CIRS: immune-inflammatory response system / compensatory immunoregulatory system ratio

**Figure 1.**
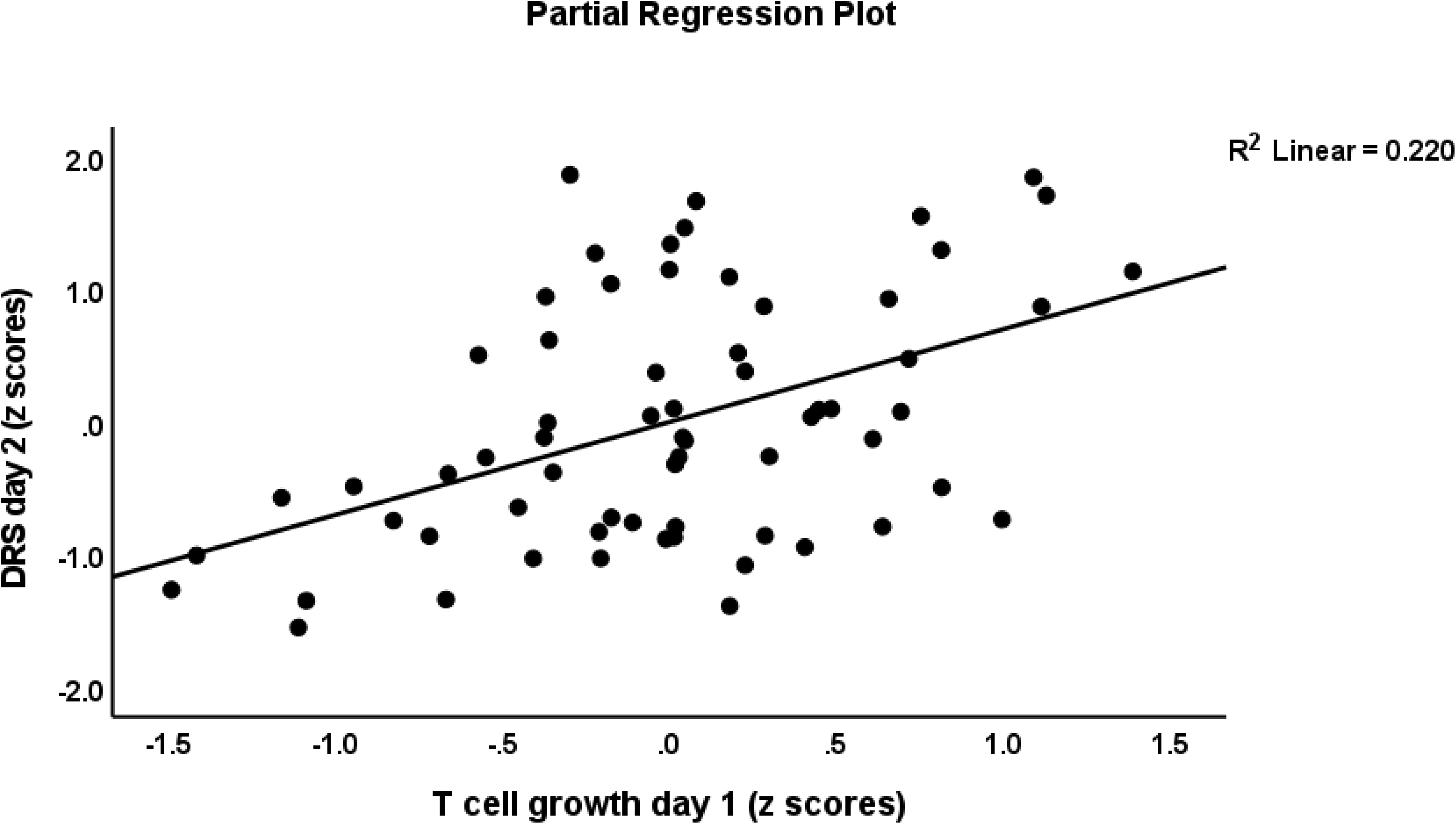
Partial regression of the Delirium Rating Scale, Revised-98-Thai (DRS) score (day 2) on the T cell growth profile measured at day 1.

**Figure 2.**
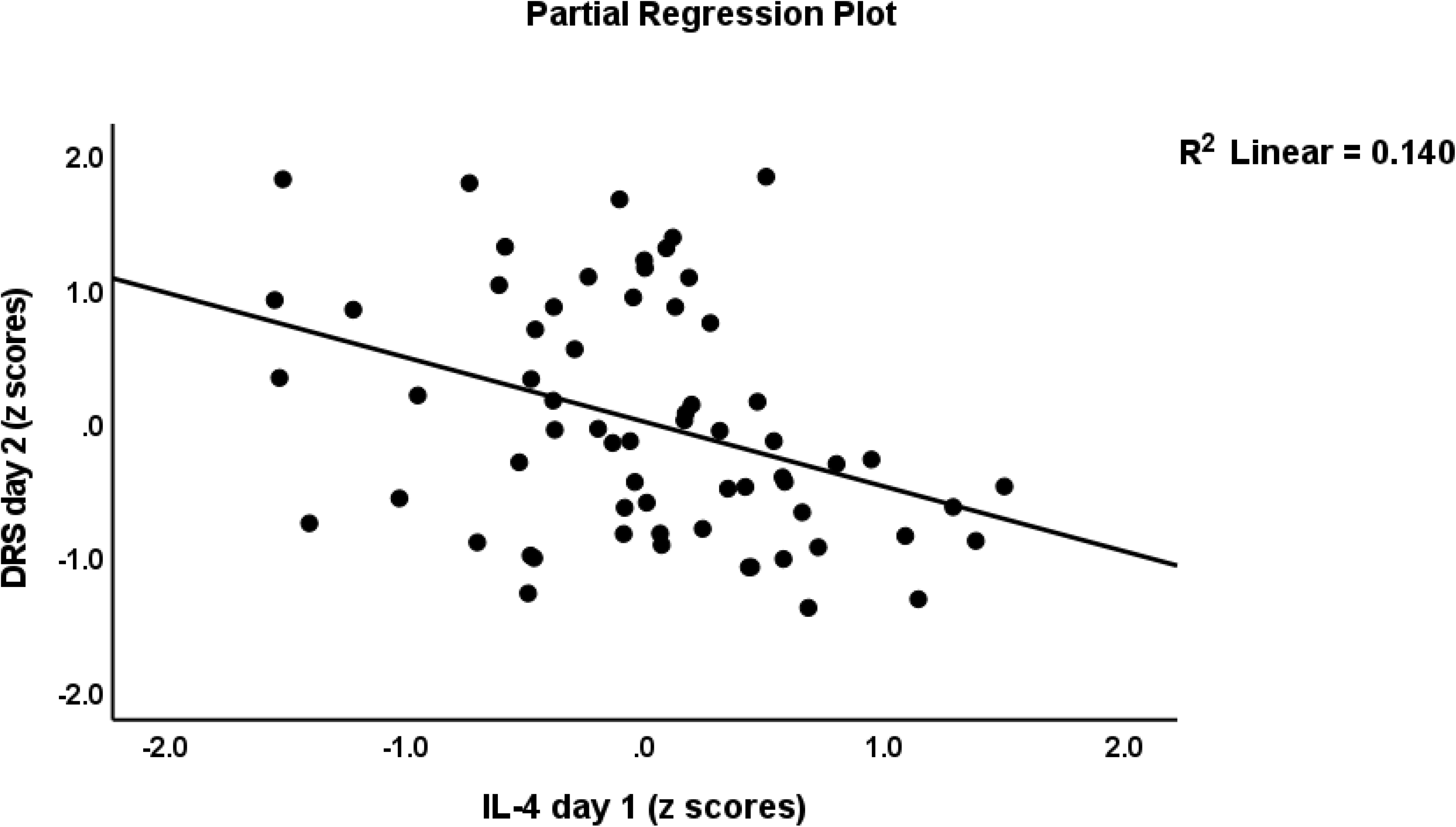
Partial regression of the Delirium Rating Scale, Revised-98-Thai (DRS) score (day 2) on interleukin (IL)-4 levels measured day 1

**Figure 3.**
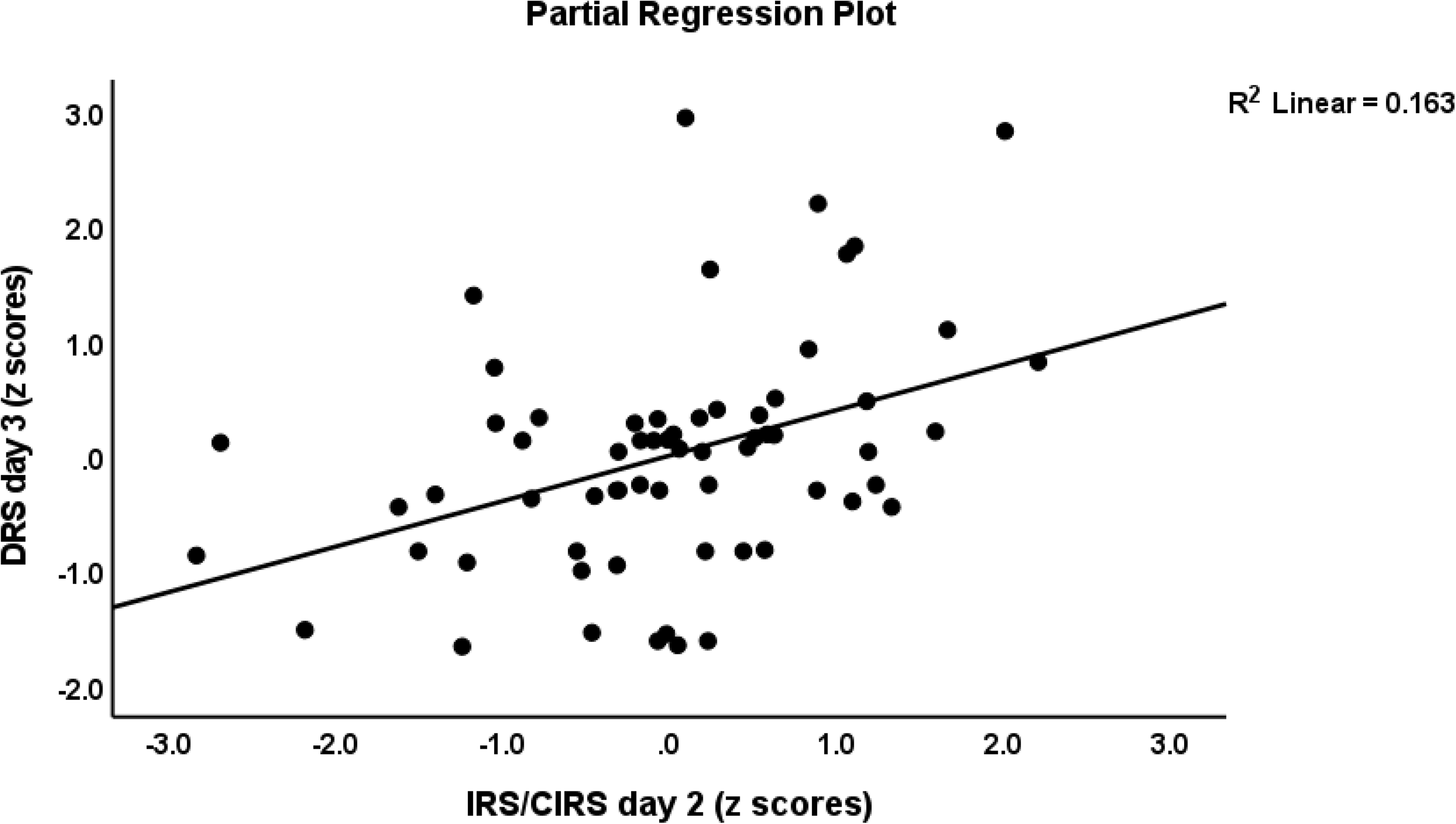
Partial regression of the Delirium Rating Scale, Revised-98-Thai (DRS) score (day 3) on the immune-inflammatory responses system (IRS) / compensatory immunoregulatory system (CIRS) ratio measured day 2

Consequently, we have delineated the immune profiles and cytokines/chemokines that predict the DRS-R-98 scores at day 1, day 2 or day 3 after adjusting for the baseline DRS-R-98 values. As such these multiple regression analyses examine the associations between the residualized DRS-R-98 scores (i.e. the actual changes from baseline to days 1. 2 or 3) and the immune profiles with and without demographic/clinical data. **Table 4** regression # 1 shows that 59.9% of the variance in DRS-R-98 score (day 1) was explained by the regression on DRS baseline, IL-8, TNF-α and T cell growth (all day 1 and positively associated) and CIRS and IL-4 (all day 1 and inversely associated). Regression # 2 shows that 38.4% of the variance in the DRS-R-98 score (day 2) was explained by the regression on the DRS baseline and Th1 (day 2) (both positively) and IL-4 (day 2, inversely associated). Regression # 3 shows that 29.7% of the variance in DRS-R-98 score at day 2 was explained by the regression on age, Th1 (day 1) and DRS baseline (all positively). Regression # 4 shows that 27.9% of the variance in the DRS-R-98 score at day 3 was predicted by the IRS/CIRS ratio (day 2) and baseline DRS (both positively associated). Regression #5 shows that 42.5% of the variance in the mean DRS-R-98 score averaged over days 1, 2 and 3 was explained by DRS baseline, age and T cell growth (day 1) (all positively). The neurotoxicity profile, NLR and HCO3-were not significant in any of the above regressions.

**Table 4.**
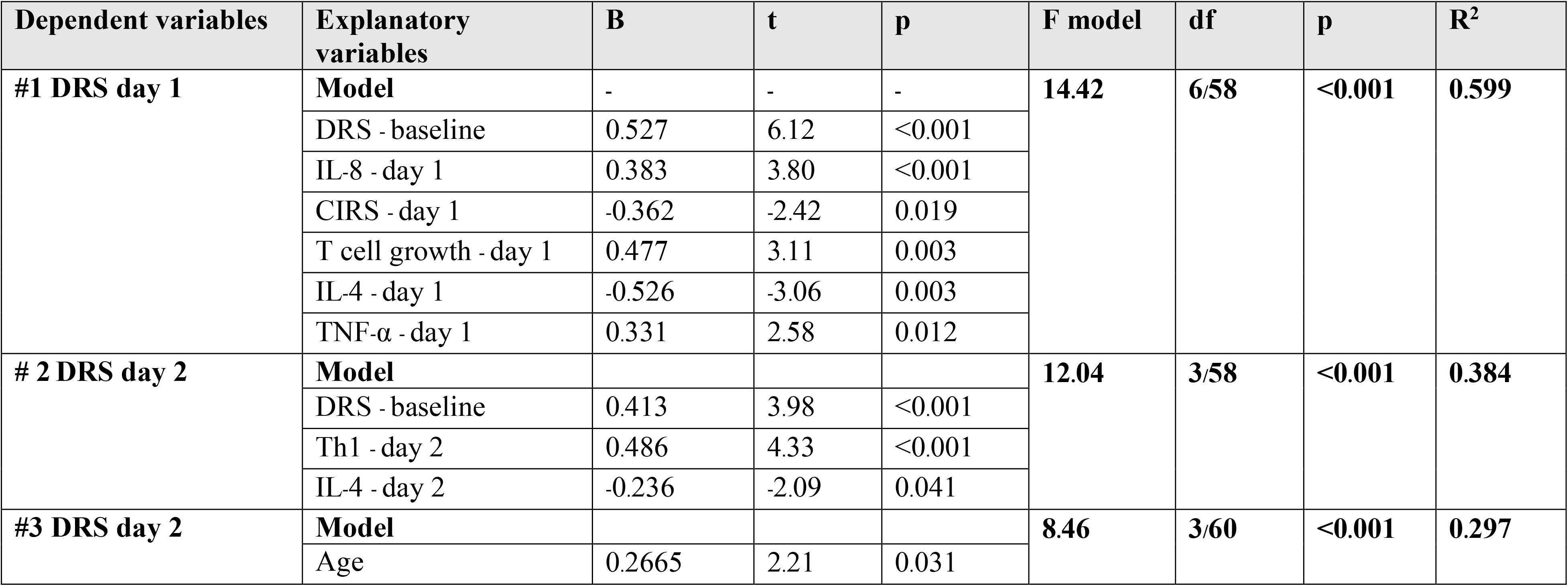
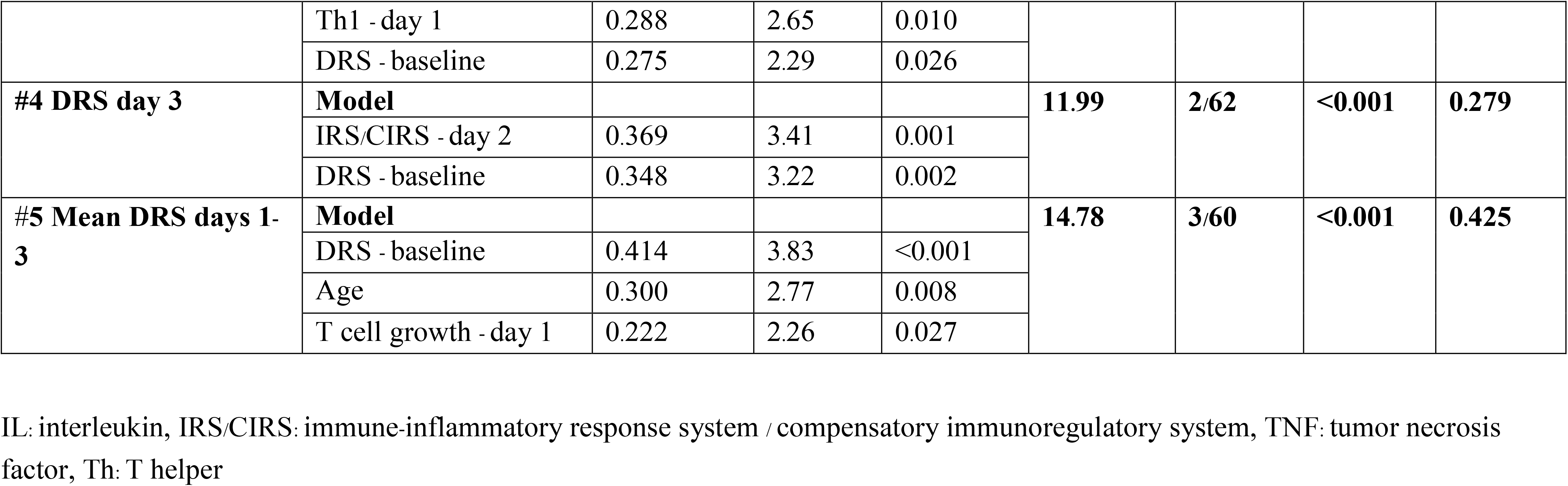
Result of multiple regression analysis with the Delirium Rating Scale, Revised-98-Thai (DRS) scores at day 1, day 2 or day 3 as dependent variables and immune/cytokine profiles and baseline DRS scores as explanatory variables

**Table 5** shows the results of multiple regression analyses with pain scores at day 1, 2 or 3 as dependent variables and cytokine profiles and clinical data as explanatory variables. Table 5, regression # 1 shows that 39.4% of the variance in the mean DRS-R-98 scores (averaged over day 1+2+ 3) was explained by HCO3-(day 1), IL-8 (day 1) and BMI (all positively) and IL-4 (day 1, inversely associated). **Figure 4** shows the partial regression of the pain scores at days 1+2+3 on the IL-8 (day 1) values. Regression # 2 shows that the pain scores (day 2) were explained by the cumulative effects of IL-8 and HCO3-(both positively, day 1) and IL-4 and sIL-1RA (day 1, both inversely associated), whereas the basal pain scores were not significant in this regression. Regression # 3 shows that 33.1% of the variance in the day 3 pain scores were explained by the regression on CCL3 (day 2), HCO3-(day 2) and BMI (all positively associated). There were no significant associations between the total DRS-R-98 score and the total pain scores (days 1, 2 and 3) (r=0.035, p=0.786, n=62).

**Table 5.** Results of multiple regression analyses with the pain scores at day 1 and 2 as dependent variables and cytokine profiles and baseline pain scores as explanatory variables

| **Dependent variables** | **Explanatory variables** | **B** | **t** | **p** | **F** | **df** | **p** | **R<sup>2</sup>** |
| --- | --- | --- | --- | --- | --- | --- | --- | --- |
| **#1 Pain, days 1+2+3** | **Model** | - | - | - | **9.26** | **4/57** | **<0.001** | **0.394** |
|  | HCO3- - day 1 | 0.447 | 4.29 | <0.001 |  |  |  |  |
|  | IL-8 - day 1 | 0.537 | 4.69 | <0.001 |  |  |  |  |
|  | IL-4 - day 1 | -0.255 | -2.27 | 0.27 |  |  |  |  |
|  | BMI | 0.221 | 2.10 | 0.040 |  |  |  |  |
| **#2 Pain, day 2** | **Model** |  |  |  | **9.97** | **4/51** | **<0.001** | **0.494** |
|  | IL-8 - day 1 | 0.699 | 5.36 | <0.001 |  |  |  |  |
|  | IL-4 - day 1 | -0.448 | -3.71 | 0.001 |  |  |  |  |
|  | sIL-1RA - day 1 | -0.386 | -3.31 | 0.002 |  |  |  |  |
|  | HCO3-- day 1 | 0.309 | 3.08 | 0.003 |  |  |  |  |
| #3 Pain, day 3 | **Model** |  |  |  | **9.55** | **3/58** | **<0.001** | **0.331** |
|  | CCL3 - day 2 | 0.471 | 4.23 | <0.001 |  |  |  |  |
|  | HCO3-- day 2 | 0.422 | 3.83 | <0.001 |  |  |  |  |
|  | BMI | 0.250 | 2.29 | 0.026 |  |  |  |  |

IL: interleukin, sIL-1RA: soluble interleukin 1 receptor antagonist, BMI: body mass index

**Figure 4.**
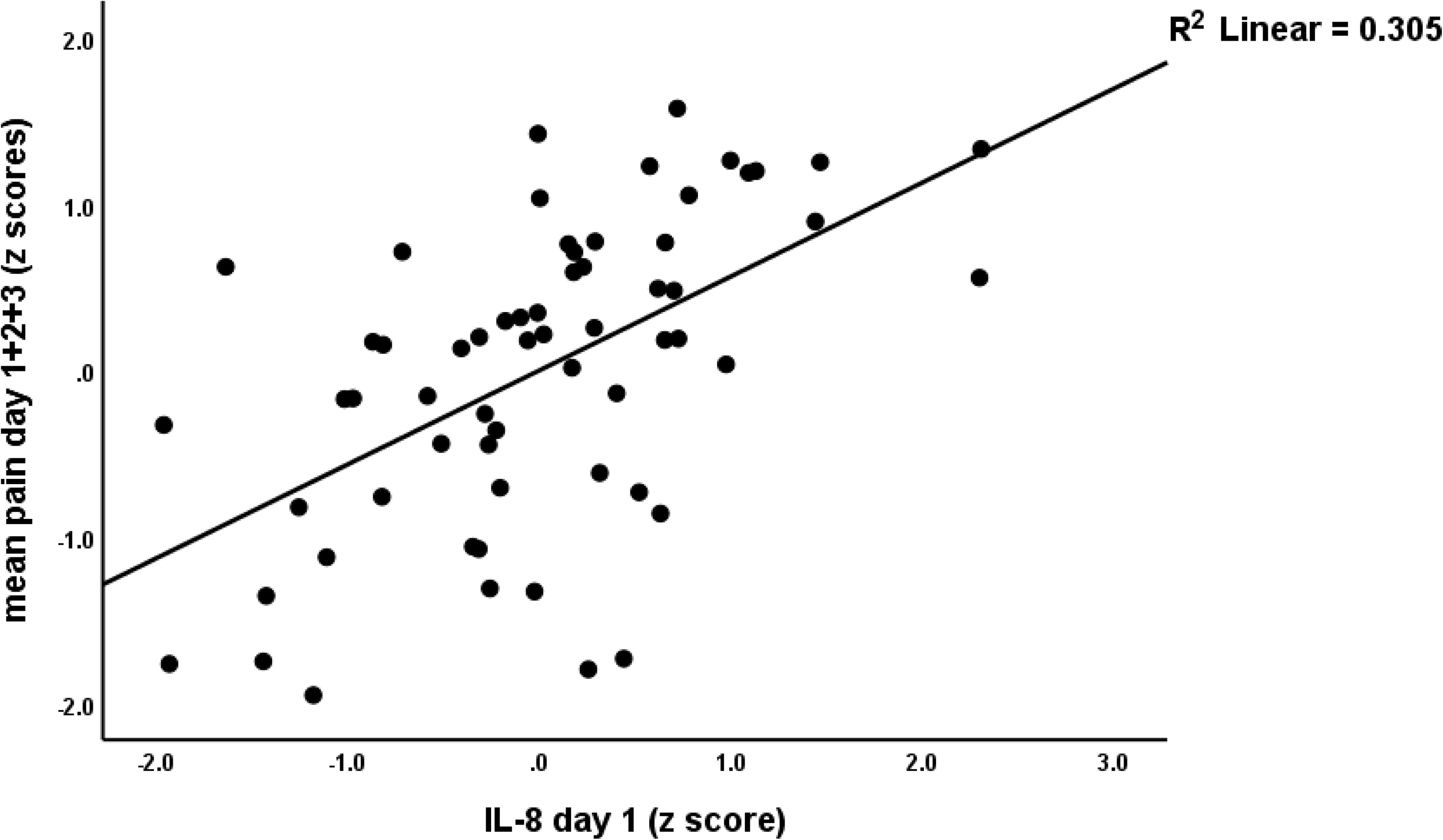
Partial regression of the pain score (mean of days 1+2+3) on interleukin (IL)-8 measured day 1

**Table 6** shows the results of GEE analyses with the NLR as dependent variable and immune profiles as explanatory variables. We found that the NLR was significantly and positively predicted by the IRS/CIRS ratio, and M1, Th1, Th17, Th2, T cell growth and neurotoxicity profiles.

**Table 6.**
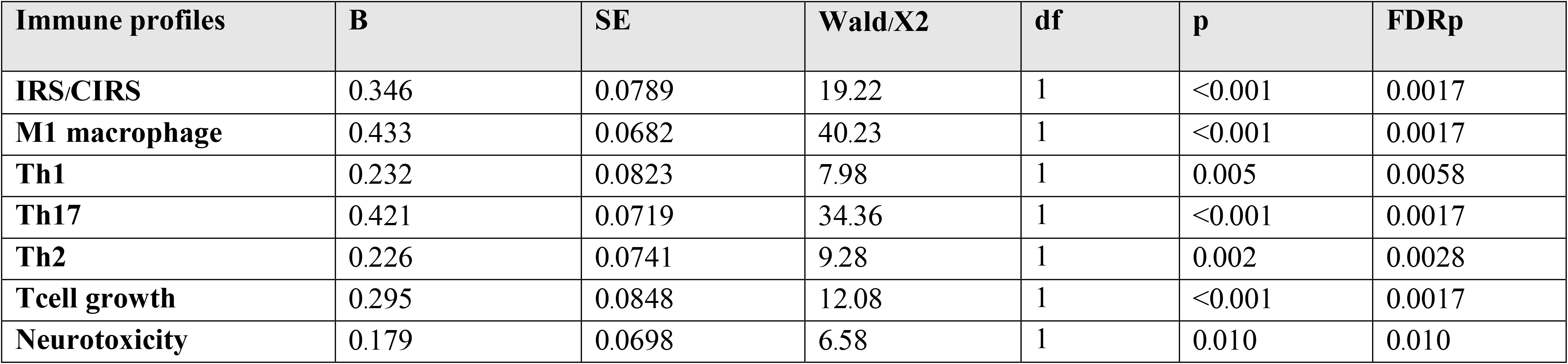
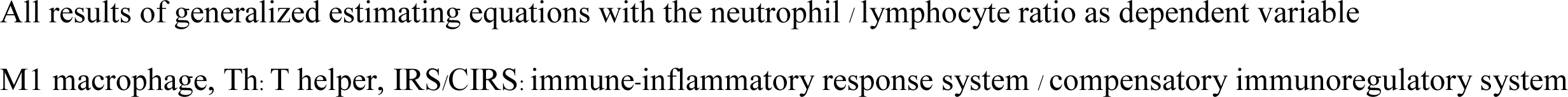
Association between the neutrophil/lymphocyte ratio and immune profiles.

| Immune profiles | B | SE | Wald/X2 | df | p | FDRp |
| --- | --- | --- | --- | --- | --- | --- |
| **IRS/CIRS** | 0.346 | 0.0789 | 19.22 | 1 | <0.001 | 0.0017 |
| **M1 macrophage** | 0.433 | 0.0682 | 40.23 | 1 | <0.001 | 0.0017 |
| **Th1** | 0.232 | 0.0823 | 7.98 | 1 | 0.005 | 0.0058 |
| **Th17** | 0.421 | 0.0719 | 34.36 | 1 | <0.001 | 0.0017 |
| **Th2** | 0.226 | 0.0741 | 9.28 | 1 | 0.002 | 0.0028 |
| **Tcell growth** | 0.295 | 0.0848 | 12.08 | 1 | <0.001 | 0.0017 |
| **Neurotoxicity** | 0.179 | 0.0698 | 6.58 | 1 | 0.010 | 0.010 |

All results of generalized estimating equations with the neutrophil / lymphocyte ratio as dependent variable
M1 macrophage, Th: T helper, IRS/CIRS: immune-inflammatory response system / compensatory immunoregulatory system

## Discussion

The first major finding of this study is that delirium and/or the severity of delirium symptoms are significantly associated with the IRS/CIRS ratio, namely positively with M1 (i.e. IL-6, CXCL8 and TNF-α), Th1 and Th17 activation and T cell growth (positively), and inversely with IL-4 and sIL-1RA. These findings extend those of previous papers showing that alterations in peripheral levels of IRS cytokines, namely IL-1β, IL-6, CXCL8, IL-10, and TNF-α, and also C-reactive protein (CRP) and NLR are associated with the onset of delirium [3, 18, 19, 39, 40].

M1-associated cytokines including IL-1β, IL-6, CXCL8, and TNF-α, play a key role in the immune response to injuries. Danger associated molecular patterns, endogenous molecules released from death cells, induce local monocytes and macrophages to secrete IL-1β [41], which is a major pro-inflammatory cytokine secreted from sterile injurious areas [42]. Increased IL-1β signaling is involved in many medical and psychiatric conditions such as tissue damage [43], sepsis [44], rheumatoid arthritis [45], as well as schizophrenia [24], and mood disorders [46]. IL-6 produced by locally damaged tissue and macrophages pleiotropically regulates CD4+ T cell differentiation including Th17 proliferation and Treg inhibition [47]. At the local injurious site, macrophages also secrete CXCL8 to promote further local and systemic inflammatory processes including neutrophil stimulation [48]. Accumulation and migration of mononuclear and polymorphonuclear cells (such as neutrophils, macrophages, lymphocytes, monocytes) to the fracture site are observed after a traumatic injury event [49]. Consequently, these immune cells and the injured tissues secrete several pro-inflammatory cytokines and chemokines which expand the inflammatory/anti-inflammatory processes from the local to the systemic level and then to the brain [50]. In this respect, TNF-α is one of these potent pro-inflammatory markers which orchestrate acute inflammatory cascades throughout the body and the central nervous system as well [51, 52]. It is important to note that the above inflammatory markers are all consistently associated with delirium [53, 54]. [39, 59-61]. Unsurprisingly, our study also observed significantly increased levels the Th1 (combination of IL-2, IFN-γ and IL-12) and T cell growth and activation factors (a combination of IL-4, IL-7, IL-9, IL-12, IL-15, GM-CSF) in the delirious hip fracture patients compared to the non-delirious group. As such, activated cell-mediated immune pathways are associated with the severity of delirium symptoms in elderly subject with post-surgery hip fracture.

Based on the above results we may conclude that delirium is accompanied by a cascade of early inflammatory mechanisms which extend from local tissue injury to inflammatory cell activation to cytokine release with increased CRP production and an increased NLR, and neuroinflammation [17]. In response to the acute phase of trauma and inflammation, locally and systematically increased IL-6 secreted from macrophages and T-cells signal the hepatocytes to produce positive acute phase proteins including CRP [55, 56]. Here we report that the NLR is a highly significant biomarker of delirium which strongly reflects IRS, M1, Th2, Th17, and IRS/CIRS activation and Tcell growth as well. As such, the present study extends previous research that high serum and CSF CRP and an increased NLR in peripheral blood are consistently reported as biomarkers or predictive risk factors of post-operative delirium [17][19, 57-60]. Nevertheless, the immune profiles assayed here better predict delirium symptoms than NLR.

IL-17 is another inflammatory mediator that is produced by CD4+ Th17 cells stimulated by IL-6 and IL-1β and by CD8 T cells and neutrophils. IL-17 plays a key role in chronic inflammatory disorders and autoimmunity, and this cytokine stimulates chemokines (e.g., CXCL1, CXCL2 and CXCL8) and granulopoiesis [61]. Due to its neurotoxic effects, increased IL-17 levels play a role in first episode psychosis and schizophrenia [62, 63], and mood [64], and neurodegenerative disorders [65].

Importantly, CXCL8 was significantly associated with the severity of delirium as assessed with the DRS-R-98 scale. Increased levels of peripheral IL-8 are reported in many neuropsychiatric disorders such as major depression, schizophrenia, bipolar disorder, autistic spectrum disorder, and Alzheimer’s disease [66-68]. In response to intracerebral pro-inflammatory stimuli, IL-8 is released from microglia and consequently attracts neutrophils and leucocytes to expand the neuro-inflammation, which may lead to a poorer CNS outcome after a traumatic brain injury or bacterial meningitis [69]. Although increased IL-15 levels in peripheral blood and the CNS play a crucial role in neuro-inflammatory disorders including multiple sclerosis, encephalomyelitis, and intracerebral hemorrhage [70-73], this study could not find any increases in this cytokine in delirium. A study in 2009 found that high IFN-γ was significantly associated with more severe delirium [74] and IFN-γ participates in the macrophage stimulating process during acute inflammation [73, 75].

The second major finding of this study is that the CIRS index and IL-4 and sIL-RA, which display anti-inflammatory effects, are inversely associated with delirium and/or the severity of delirium symptoms. The sIL-RA is secreted by activated macrophages and inhibits pro-inflammatory IL-1β signaling [76] and, therefore, the decrease in sIL-RA levels as seen the current and previous studies [74, 77], suggests lowered negative feedback on pro-inflammatory IL-1 signaling [78]. IL-4, which is secreted by Th2 lymphocytes, basophils, mast cells, and eosinophils, leads to the modification and proliferation of lymphocytes, macrophages, fibroblasts and endothelial cells and promotes anti-inflammatory and immunoregulatory processes, as well as healing of tissues [79]. Interestingly, IL-4 function is associated with neuro-restorative effects after cerebral ischemic and traumatic brain injury events, while lower IL-4 signaling is associated with cognitive impairments in schizophrenia [80]. Moreover, relative lower CIRS activity in first episode psychosis predicts a worse outcome following treatment with antipsychotics [81]. Previous research showed that the anti-inflammatory cytokine IL-10, which is produced by different cells but especially by Treg cells, may regulate the initial pro-inflammatory response in delirium [82, 83]. Overall, lowered anti-inflammatory defenses through lowered CIRS functions appear to contribute to delirium.

The third major finding of this study is that the neurotoxic immune profile comprising neurotoxic cytokines and chemokines was not associated with delirium. This contrasts with the increased neurotoxic profiles established in schizophrenia, cognitive impairments in schizophrenia, mood disorders, and suicide attempts and ideation [84-86]. At the phenomenological level, post-hip fracture operative delirium has an acute onset and is frequently presenting with more positive psychotic features [87] contrasting with the chronic nature of schizophrenia and mood disorders. Thus, the acute increase in IRS (M1, Th1 and Th17) cytokines in patients with attenuated CIRS functions appears to be the most important factor in delirium.

It is important to note that the increased cytokine levels produced at the traumatic site or peripheral vascular and lymphatic system may pass through a damaged blood brain barrier or circumventricular organs to signal inflammatory cascades in the central nervous system [88, 89]. For example, significantly elevated levels of peripheral cytokines including IL-1β, IL-6, IL-8, TNF-α, and IFN-? are involved in the peripheral to central inflammatory signal transduction [52, 90]. Peripheral inflammatory processes which translate peripheral inflammatory signals into central neuro-inflammation and microglial activation are described in delirium [18] and unipolar depression [91] and psychosis [63].

The fourth major finding of this study is that higher pain scores in post-surgery patients are positively associated with CXCL8 and CCL3, and inversely with IL-4 and sIL-1RA. The pain scores were not related to delirium or severity of delirium which contrasts with previous studies which identified increased pain scores as a major contributing factor to postoperative delirium [92, 93]. Almost all of the patients in our cohort received an ultrasound-guided fascia iliaca block pre-operatively [94, 95] which may contribute to attenuated peri-operative pain experiences. Similar to our findings, increased CXCL8 and CCL3 levels, and decreased IL-4 and IL-1RA level were previously reported to be associated with neuropathic pain [34].

The results of the present study should be discussed with regard to its limitations. A first limitation is that we did not distinguish between delirium subtypes including the hyperactive, hypoactive, and mixed phenotypes. The different delirium subtypes may have a different pathogenesis and may represent different immune-inflammatory and neurochemical pathways [96]. Second, the biomarkers analyzed in this study reflect the peripheral part of the IRS response and not the neuro-inflammatory changes in the CNS which should be examined by cerebrospinal fluid analyses or brain imaging techniques [97].

## Conclusion

Delirium, the severity of delirium and the changes in the DRS-R-98 score from baseline to post-surgery are associated with IRS activation as indicated by M1, Th1, Th17 and T cell growth profiles. The latter changes predict delirium, especially when CIRS functions including IL-4 and sIL-1RA levels are attenuated. The development of delirium is not associated with a neurotoxic cytokine/chemokine profile as observed in mood disorders and schizophrenia, suggesting that the acute effects of IRS cytokines are sufficient to induce neurocognitive impairments, and psychomotor and psychotic symptoms. Pain ratings during the post-operative period are substantially and positively linked with CXCL8 and CCL3, but inversely with IL-4 and sIL-1RA. Increased NLR expression is indicative of overall immunological and M1/Th1/Th17/Th2/Treg cell activation.

## Data Availability

All data produced in the present study are available upon reasonable request to the authors

## Ethics statement

Approval for the study was obtained from the Institutional Review Board of the Faculty of Medicine, Chulalongkorn University, Bangkok, Thailand (registration number 528/61), in compliance with the International Guideline for Human Research protection, as required by the Declaration of Helsinki, was conducted according to Thai and international ethics and privacy laws.

## Informed consent

Written informed consent was obtained before the study from the patients or their gardians (first-degree family members).

## Authors’ contributions

All the contributing authors have participated in the preparation of the manuscript.

## Acknowledgements

We gratefully acknowledge the help of all psychiatry/orthopedic/anesthesiology nursing staff and residents involved in the execution of this study.

## Data Access Statement

The dataset generated during and/or analyzed during the current study will be available from MM upon reasonable request and once the dataset has been fully exploited by the authors.

